# Device-measured physical activity, sedentary time, and sleep in patients with arrhythmogenic cardiomyopathy: descriptive values and stability over 30 measurement days

**DOI:** 10.1101/2022.05.20.22275318

**Authors:** David Ruiz-González, Javier Ramos-Maqueda, Jairo H. Migueles, José Antonio Vargas-Hitos, Amalio Ruiz-Salas, Juan Jiménez-Jáimez, Alberto Soriano-Maldonado

## Abstract

**Introduction:** Regular exercise and sports are contraindicated in arrhythmogenic cardiomyopathy (AC) patients, which might lead them to an unhealthy sedentary lifestyle. This study aimed to objectively describe the patterns of physical activity (PA), sedentary time (SED), and sleep in patients with AC, and to examine the reproducibility of accelerometer-derived measures over four consecutive weeks.

**Methods:** A total of 71 (49.6 [SD=17.5] years) patients with AC wore a wrist-worn Axivity AX3 accelerometer for 30 consecutive days to monitor their physical activity, sedentary time, and sleep habits. The reproducibility of each metric across the 4 assessment weeks was assessed with the intraclass correlation coefficients (ICCs) derived from linear-mixed models adjusted for age, body mass index (BMI), and season.

**Results:** The participants spent a median of 12.2 [IQR 2.1] h/d in SED, 6.4 [IQR 1.0] h/d sleeping, and 17.9 [IQR 24.5] min/d in moderate-to-vigorous physical activity (MVPA), and 59% of the participants did not reach the 150 min/d of MVPA recommended by the WHO for people living with chronic disease. No significant differences in PA were found by sex and age groups. Otherwise, patients aged ≥50 years (n= 33) spent 38.9 min/d (95% CI 5.8 to 72.2, p≤0.05) more in periods of ≥30 minutes of SED than those <50 years. Participants with obesity (n=10) accumulated 66.6 min/d (95% CI 5.2 to 128.1, p = <0.05) more SED in periods of ≥30 minutes and 22.8 min/d (95% CI 0.7 to 44.9, p≤0.05) less MVPA than those without obesity. The ICCs ranged from 0.67 for time in bed to 0.92 for light-intensity physical activity using a 7-day assessment period. However, the ICCs increased from 0.03 for LPA to 0.18 for time in bed when an assessment period of 14 days was chosen.

**Conclusion:** Patients with AC engage in large periods of SED, insufficient PA and sleep. Importantly, nearly 60% of the participants did not meet the minimum amount of PA recommended by the WHO for people living with chronic diseases and only 20% met the sleep recommendations. Device-measured PA and SED are stable across weeks, indicating that a 7-day assessment period might provide a reproducible measure of PA and SED and, to a lower extent, sleep.

## 1. Introduction

Arrhythmogenic cardiomyopathy (AC) may lead to ventricular arrhythmias and sudden cardiac death.^1^ The AC estimated prevalence ranges from 1:1000 to 1:5000, and 0.08% to 3.6% of the patients die annually.^2^ AC is a genetic disease of the cardiac muscle characterized by loss of cardiomyocytes and their replacement by fibrous or fibrofatty tissue. Dysfunctional desmosomes play a key role in the pathogenesis of AC,^3^ resulting in a progressive myocyte death and reduced wall stress tolerance. This leads to an early cardiac remodeling of the right ventricle with potential involvement of the left ventricle.^4^ AC usually onsets in the adulthood, with a median age at diagnosis of 31 years and a greater prevalence in males.^5–7^

The available evidence indicates that decedents with a diagnosis of AC are more likely to die at rest or during sleep. This might be related to the fact that regular exercise and competitive sports are contraindicated in AC patients.^8–11^ In fact, the European Society of Cardiology guidelines recommend that individuals with AC must avoid participation in high-intensity exercise because they have been related with accelerated disease progression, greater risk of sudden cardiac arrest and death.^12,13^ Nevertheless, recent World Health Organization (WHO) guidelines for adults^14^ highlights the importance of not depriving individuals with chronic conditions from the many benefits of physical activity (PA), and the need to safely participating in activities of low to moderate intensity.^15–18^ Similarly, engaging in large periods of sedentary time (SED) is a widely recognized independent risk factor for cardiovascular morbi-mortality,^19^ and insufficient sleep is also associated with cardiovascular events including, arrhythmia, atrial fibrillation, and myocardial infarction.^20–22^ Understanding how AC engage in health-related behaviors such as lifestyle PA, SED or sleep it is of wide clinical relevance because they are strongly associated with all-cause and cardiovascular morbidity and mortality in many populations^23^ including patients with cardiovascular disease^24^ and arrythmia.^25^

Previous research on AC patients have focused on self-reported regular exercise habits and/or sports participation (either competitive or recreational).^12,13,26–28^ To date, the lifestyle activity patterns of people living with AC have not been described. Activity monitors (e.g., accelerometers) are useful to non-invasively assess the PA, SED, and sleep habits, overcoming some of the bias that affect self-reported data (e.g., social desirability or recall bias).^29–33^ Characterizing PA, SED and sleep in AC patients using objective methods is relevant to guide future clinical guidelines regarding these important health-related behaviors. In addition, although the standard accelerometer data collection protocols last approximately seven days, the week-to-week stability of the device-based measures has been questioned.^34^ The only study investigating the week-to-week stability of the device-measured PA found stable and reproducible estimates in older women.^34^ Assessing the week-to-week stability of PA, SED, and sleep using accelerometers in AC patients is of clinical interest to understand what is the most recommended measurement period that provides reproducible estimates with the minimal amount of resources. Therefore, the aims of this study were (i) to provide a detailed description of PA intensity parameters (light physical activity (LPA) and moderate-to-vigorous physical activity (MVPA)), SED, and sleep in a sample of Spanish adults living with AC; and (ii) to quantify the week-to-week stability of the PA intensities, SED, and sleep over a period of 30 days.

## 2. Methods

### 2.1. Study design and participants

We performed an observational study involving a total of 72 patients with AC. Patients were recruited from the clinical cohorts of two tertiary hospitals with units specialized in the care of inherited heart diseases: the Virgen de las Nieves University Hospital (Granada, Spain) and the Virgen de la Victoria University Hospital (Malaga, Spain). Patients included were required to be diagnosed with AC as determined by the revised 2010 Task Force Diagnostic Criteria^35^ or patients with a causative mutation for AC without the requirement to fulfil the criteria established by the 2010 task force. Additional inclusion criteria were the presence of a definitive diagnosis of left-dominant AC according to Padua Criteria.^37^ Of the 72 AC patients initially recruited, patients without accelerometer data were excluded from analyses (n=1), using a sample of 71 patients.

The Research Ethics Committee of Granada approved the study protocol, and the research was conducted in accordance with the principles of the Declaration of Helsinki. Written informed consent was obtained from all participants.

### 2.2. Clinical Data Acquisition

Sociodemographic characteristics, medical history, and device-measured PA, SED, and sleep were assessed. During this same period, arrhythmia measurement was performed by Nuubo textile Holter^®^, a three-channel electrocardiogram recording system with cardiac event detection algorithms.^38^ Moreover, all patients were genetically characterized using large AC Next-Generation Sequencing panels in probands and Sanger sequencing cascade screening in relatives, and information regarding previous serious arrhythmic events, such as cardiac arrest, sustained ventricular arrhythmias or appropriate therapy of the Implantable cardioverter-defibrillator (ICD), was collected.

### 2.3. Accelerometer data collection and processing

To measure PA, SED and sleep parameters patients wore a triaxial accelerometer (Axivity AX3©, Axivity Ltd, United Kingdom). The AX3 has been validated and used for measuring PA in a previous large-scale cohort study.^39^ Patients were instructed to wear the accelerometer on the non-dominant wrist 24 hours per day, for 30 consecutive days. A valid measurement day was considered when participants wore the accelerometer for at least 16 hours per day. For a valid measurement week, participants must have recorded at least four valid days.

The accelerometer was set at a sampling frequency of 25 Hz, which is enough to capture most of the daily life accelerations related to physical movement and provides a close measurement to the standard 100 Hz used in other studies.^40^ This decision was made (instead of the standard protocols with higher sampling frequencies) to ensure the battery life and storage capacity of the device for the 30 days of data collection. Raw accelerometer data files were processed with the R GGIR software (v. 2.5-0, https://cran.r-project.org/web/packages/GGIR/).^41^ In brief, the GGIR pipeline included: (i) auto-calibration of the raw accelerations to the local gravity; (ii) aggregation of the acceleration signal over 5-s epochs after removing the gravitational acceleration;^42,43^ (iii) non-wear time detection using 15-minute blocks with a 60-minute sliding window when the standard deviation of two out of the three axes was lower than 13 m*g* or the value range in two of the three axes was lower than 50 m*g*;^44^ (iv) imputation of the non-wear time detected; (v) classification of the sleep and awake periods using automated algorithms;^45,46^ (vi) classification of the awake time into SED or physical activity of light, moderate, or vigorous intensity.^47,48^

Sedentary time (<40 m*g*), and physical activity of light (LPA, 40-100 m*g*) and moderate-to-vigorous intensity (MVPA, ≥100 m*g*) were classified based on previously-validated acceleration thresholds.^47,48^ MVPA periods lasting less than one minute (for which 80% of the acceleration were over 100 m*g*) were considered of light intensity as they likely represent random wrist movement.^49^ Additionally, sedentary bouts lasting more than 30 consecutive minutes were identified with a tolerance of 10% of the bout time. Total sleep duration (minutes classified as asleep within the sleep period time (or time in bed)), time in bed, and sleep efficiency (the proportion of time spent sleeping from onset to termination: (sleep duration–wake after sleep onset (WASO)) / time in bed). Sleep efficiency ranges from 0 to 100, where a score of 100 indicates that the individual did not wake up between sleep onset and termination.

### 2.4. Statistical analysis

For descriptive analysis of metrics related to sleep, SED and PA, participants were grouped on sex and age (<50 years, ≥50 years) that were self-reported. Height and weight were measured, and BMI was calculated as weight (kg) divided by squared height (m^2^). Overweight and obesity were defined as BMI 25-29.9 and ≥30 kg/m^2^, respectively, and three age groups were created (normal weight, overweight and obesity). In addition, differences in PA levels were analyzed for disease-related variables that might affect the amount of PA. These variables divided participants according to ICD and the presence of desmosomal protein gene mutations.

Due to the non-normal distribution of most PA variables (assessed with the Kolmogorov Smirnov test), descriptive characteristics are presented using medians and interquartile ranges (IQR). Differences among groups stratified by socio-demographic and clinical characteristics in normally distributed numerical variables were tested using one-way analysis of variance (ANOVA), and by Mann-Whitney U test in case of deviation from normal distribution. For each activity metric, the weekly average was calculated and Friedman test with Bonferroni’s adjustment were used to estimate whether the accelerometer metric changed over time. To assess the stability over the four weeks for all participants combined, the intraclass correlation coefficients (ICC) was used by dividing the between-person variance by the sum of the between-person and within-person variances. Between-person and within-period variances were estimated using random effects models that included average weekly value for each metric at each of the four weeks. ICCs are presented with and without adjustment for age, BMI, and season of the year. Ninety-five percent confidence intervals (95% CI) were calculated for the ICCs based on a two-way mixed effects model, using single measures, and absolute agreement. As there were six subjects who had no accelerometer data for all four weeks, a sensitivity analysis was conducted to examine the impact of the different sample sizes at weeks two, three and four on the ICCs. Further, reliability associations were classified based on the 95% confidence intervals of the ICC estimate as poor (0–0.49), moderate (0.50–0.74), good (0.75–0.89), or excellent (0.90–1.00).^50^ In order to avoid potential measurement reactivity, ICCs were also calculated by eliminating the first day of measurement. To assess the utility of a single 7-day measurement, Spearman’s Rank Order Correlation test was used to test the relationship between the values obtained in each of the four weeks of measurement. All statistical analyses were performed using SPSS version 25.0 (SPSS Inc., Chicago, IL, USA) and Stata (version 16.1.0; Stata Corporation, College Station, TX, United States). Values of p < 0.05 were considered statistically significant.

## 3. Results

Characteristics of the study population are presented in **Table 1**. Of the overall study participants (n = 71), median age was 51.5 (IQR 24.0) years (range from 16 to 78 years) and 36/67 (50.7%) were women. 52.1% had an ICD and 28.8% were symptomatic based on New York Heart Association functional classification (NYHA) class (>1). A total of 34.8% had documented sustained ventricular tachycardia or ventricular fibrillation and 23.1% a history of syncope. Most of the patients were carriers of a pathogenic mutation (including probands and cascade-screening detected relatives), most of them in non desmosomal genes (Table 1).

**Table 1.**
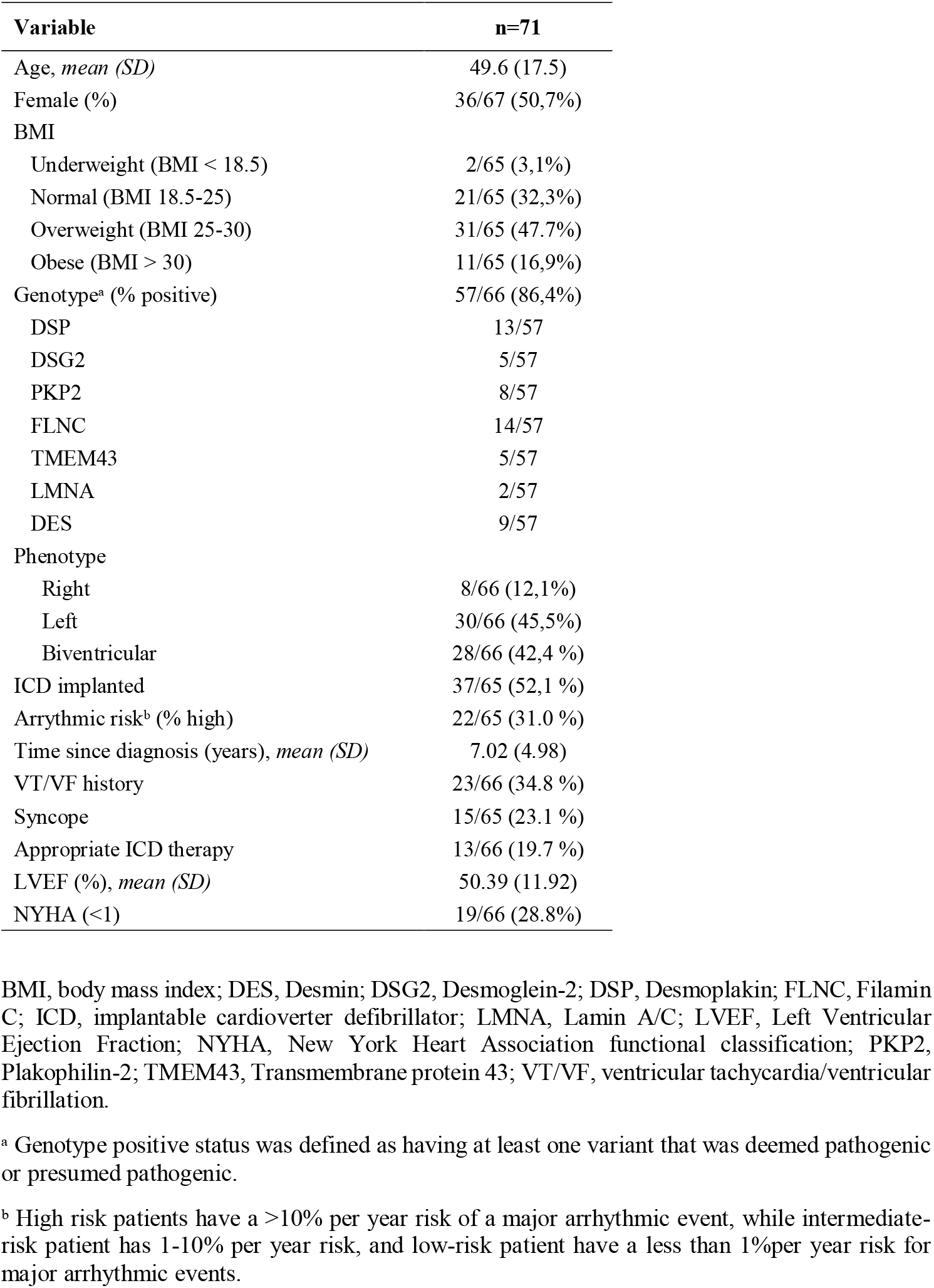
Baseline characteristics

### 3.1. Physical activity, sedentary time, and sleep

Participants had a median (IQR) of 21.0 (1.0) valid weekdays and 8.0 (0) valid weekend days, and non-wear time during the awake time was 0.9% (3.1). Accelerometer data were obtained from 71 AC individuals, of whom 65 had data from four consecutive weeks and satisfied requirements for a minimum of 2/3 day wear time on a minimum of 4 days a week. Overall, participants spent 12.2 [IQR 2.1] h/d in SED and only 17.9 [IQR 24.5] min/d in MVPA.

There were small differences in PA and SED by sex and age (Table 2). Participants ≥50 years spent a greater amount of time is accumulated in periods of ≥30 minutes of SED (94.6 min/d [IQR 102.9]) in comparison to those <50 years (50.4 min/d [IQR 49.1]). Similarly, for BMI groups, obese participants had twice as much time in periods of at least 30 minutes of SED as normal weight participants (95.3 min/d [IQR 160.7] vs. 45.8 min/d [IQR 42.3]), in contrast to the time spent in MVPA, with lower levels compared to the normal weight group (8.3 min/d [IQR 16.3] vs. 20.8 min/d [IQR 22.9]). Overall, 59.2% of participants did not reach the 150 min/d of MVPA recommended by the WHO based on accelerometer data. Regarding sleep parameters, the sleep efficiency was 90.0%, with a mean of 96.2 min/d of WASO. There were no marked differences in these sleep patterns according to age, sex, and BMI groups. Nevertheless, when considering clinical characteristics, differences in sleep-related variables did emerge. Participants with ICD showed a reduction in sleep time (6.2 h/d [IQR 0.8] vs. 6.6 h/d [IQR 1.4]) as well as efficiency (88.3 % [IQR 4.0] vs. 90.8 % [IQR 4.8]). Similarly, patients with desmosomal mutations showed a reduction in both sleep time and efficiency. Based on the recommendation of a minimum of 7 hours of sleep for adults to promote optimal health, 80% of participants do not meet the sleep guidelines.

**Table 2:** Time spent in sleep, sedentary behavior, light physical activity, and moderate-to-vigorous physical activity in the whole sample and stratified by sex and age.

|  |  | SLEEP |  |  | Sedentary |  | LPA | MVPA | Guidelines |
| --- | --- | --- | --- | --- | --- | --- | --- | --- | --- |
|  | *n* | Time in bed (h/d) | Total sleep (h/d) | Efficiency (%) | SEDTotal (h/d) | SED30min (min/d) | Total (h/d) | Total (min/d) | Total<sup>a</sup> |
| All | 71 | 7.14 (0.94) | 6.36 (0.97) | 90.10 (5.14) | 12.16 (2.11) | 61.30 (82.14) | 3.41 (1.68) | 17.94 (24.50) | 29 (40.8%) |
| Men | 36 | 7.09 (0.84) | 6.26 (0.82) | 88.66 (5.27) | 12.38 (1.63) | 65.61 (93.23) | 3.39 (1.56) | 18.84 (27.91) | 17/28 (60.7%) |
| Women | 29 | 7.14 (1.30) | 6.36 (1.32) | 90.18 (5.27) | 11.85 (2.44) | 57.73 (80.69) | 3.73 (1.86) | 17.87 (25.00) | 11/28 (39.3%) |
| **p** |  | 0.428 | 0.271 | 0.376 | 0.142 | 0.704 | 0.214 | 0.894 |  |
| <50 | 29 | 7.16 (1.18) | 6.36 (0.75) | 88.51 (6.32) | 12.28 (3.67) | 50.36 (49.14) | 3.41 (2.04) | 19.31 (24.70) | 15/26 (57.7%) |
| ≥50 | 33 | 7.13 (1.15) | 6.28 (1.10) | 90.25 (4.79) | 12.14 (1.47) | 94.59 (102.93) | 3.47 (1.62) | 13.51 (28.81) | 11/26 (42.3%) |
| **p** |  | 0.856 | 0.752 | 0.350 | 0.357 | **0.002** | 0.209 | 0.386 |  |
| Normal | 21 | 7.33 (1.06) | 6.52 (1.13) | 90.39 (4.51) | 11.62 (3.33) | 45.75 (42.75) | 3.73 (1.80) | 20.77 (22.94) | 11/24 (45.8%) |
| Overweight | 27 | 7.13 (0.89) | 6.27 (0.96) | 88.54 (4.53) | 12.29 (1.38) | 84.13 (77.89) | 3.43 (1.82) | 15.43 (28.69) | 11/24 (45.8%) |
| Obesity | 10 | 7.05 (1.62) | 6.30 (1.22) | 90.20 (5.58) | 12.07 (2.71) | 95.27 (160.68) | 3.45 (1.75) | 8.33 (16.29) | 2/24 (8.3%) |
| **p** |  | 0.856 | 0.169 | 0.083 | 0.342 | **0.025** | 0.975 | **0.047** |  |
| N-ICD | 26 | 7.26 (1.19) | 6.64 (1.37) | 90.81 (4.79) | 11.74 (2.52) | 59.51 (91.99) | 3.10 (1.96) | 18.82 (23.78) | 8/25 (32.0%) |
| ICD | 37 | 6.96 (0.97) | 6.22 (0.75) | 88.28 (4.01) | 12.34 (1.82) | 60.00 (88.26) | 3.68 (1.54) | 15.85 (27.59) | 17/25 (68.0%) |
| **p** |  | 0.126 | **0.013** | **0.008** | 0.094 | 0.239 | 0.476 | 0.359 |  |
| N-DM | 30 | 7.22 (0.99) | 6.49 (0.88) | 90.50 (4.81) | 12.38 (1.93) | 54.60 (68.86) | 3.39 (1.80) | 20.51 (23.39) | 16/27 (59.3%) |
| DM | 32 | 6.98 (0.93) | 6.14 (0.99) | 87.93 (3.74) | 11.91 (2.25) | 83.18 (130.04) | 3.70 (1.68) | 11.96 (26.92) | 11/27 (40.7%) |
| **p** |  | 0.578 | 0.081 | **0.032** | 0.546 | 0.248 | 0.510 | 0.186 |  |

Data are medians and interquartile ranges. P-values denote significance of non-parametric test for trend across categories ( $P < 0.05$ ). Values are rounded. DM = Desmosomal mutation; ICD = Implantable cardioverter-defibrillator; LPA = light-intensity physical activity; MVPA = moderate-to-vigorous physical activity.
<sup>a</sup> Participants meeting physical activity guidelines (%) of 150 min or more moderate–vigorous physical activity per week.

### 3.2. Stability of Accelerometer-Assessed PA, SED and Sleep

The unadjusted ICCs (95% CIs) ranged from 0.60 (95% CI=0.49, 0.71) for time in bed to 0.93 (95% CI=0.90, 0.95) for LPA when a 7-day assessment period was considered, however, 14-day administration of accelerometers resulted in better ICCs and ranged from 0.77 (95% CI=0.65, 0.85) for time in bed to 0.95 (95% CI=0.92, 0.97) for LPA (Table 3). Specifically, for time in bed and sleep variables, stability increased from moderate to good when a 14-day administration was used (0.60 vs. 0.77 and 0.66 vs. 0.78 respectively). ICC values were slightly higher for most variables with adjustment for age, BMI, and season (Table 3). In the stratified analysis, the ICCs were different for all activity metrics between younger and older (Table 4). Younger group (<50 years) showed higher ICCs for SED and LPA metrics. However, for MVPA, the ICCs were significantly higher for older group (0.85 and 0.75, respectively), which may be related to their lower participation in these PA intensities. Similarly, when stratified by BMI, the group composed by obese individuals showed a higher ICC for MVPA, compared with normal weight group (0.94 and 0.65, respectively). In addition, ICCs were generally higher for the winter season, with increases for time in bed compared to the spring season (0.71 and 0.58, respectively) as well as for the total amount of MVPA compared to the spring and autumn seasons (0.90, 0.69 and 0.80, respectively). A sensitivity analysis shows similar ICC values for the 65 participants with complete data at all weeks, compared with the 71 used in the main analysis who had data at baseline and week 2, week 3, week 4, or all of them (Table 5). However, for MVPA, ICCs were higher for the group of 71 that included participants with no data over the four weeks compared to the 65 with complete data across four weeks (0.97 and 0.77, respectively). The exclusion of the first day of measurement revealed similar ICCs for both the 7-day and 14-day measurement periods compared to the values obtained without excluding the first day (Table 6). Spearman’s rank order correlations between week 1 and remaining weeks indicated significant correlations for MVPA, LPA and SED, with scores above 0.8 (Supplementary Table 1).

**Table 3:**
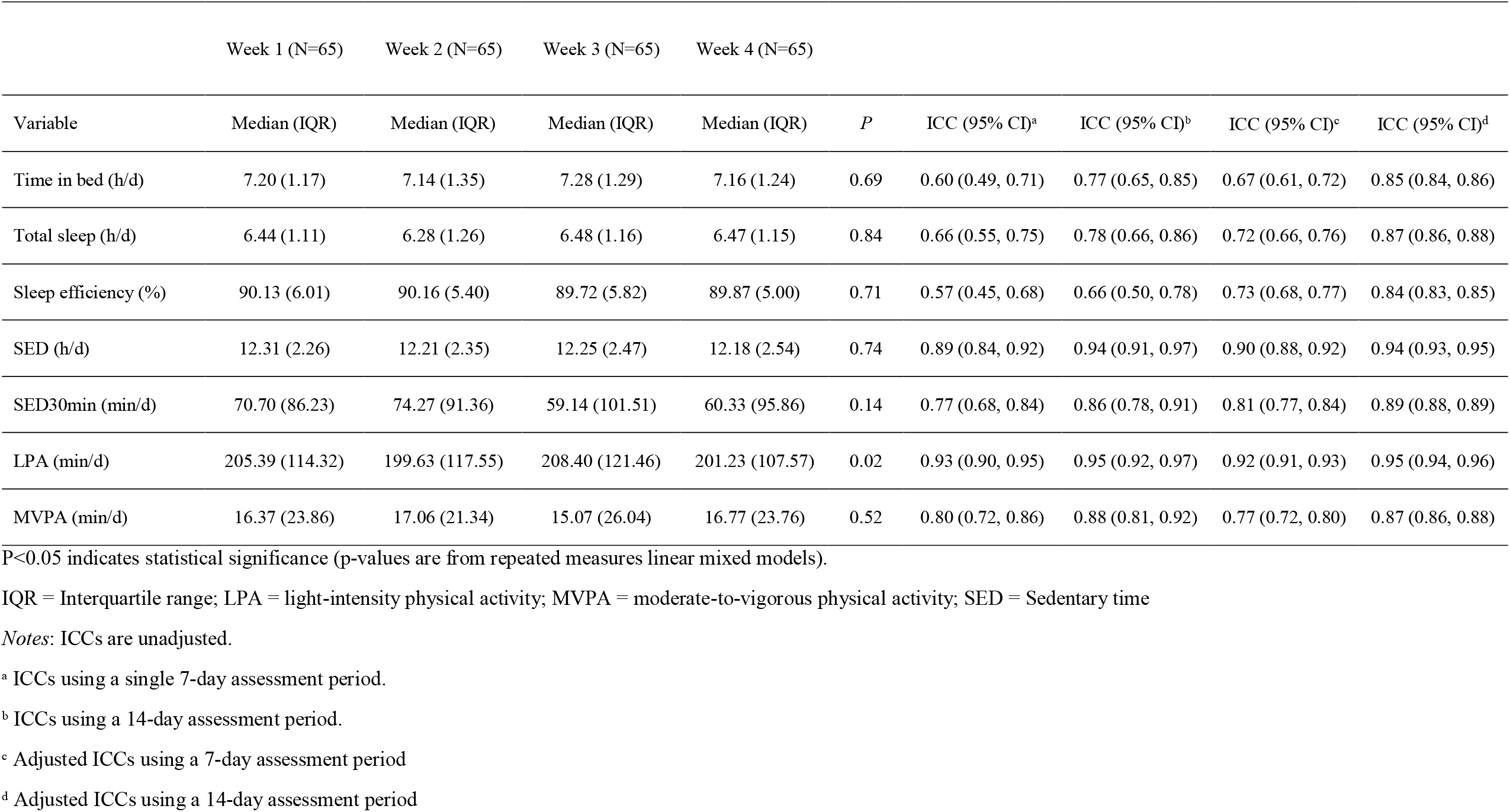
Descriptive Statistics and Intraclass Correlation Coefficient (ICC) for physical activity, sedentary time and sleep over 4 weeks.

**Table 4:** Intraclass Correlation Coefficient (ICC) for Activity Metrics stratified by Age, BMI and season over 4 weeks.

| Variables | n | Time in bed (h/d) | Total Sleep (h/d) | SED (h/d) | SED30min (min/d) | LPA (min/d) | MVPA (min/d) |
| --- | --- | --- | --- | --- | --- | --- | --- |
| Younger | 27 | 0.61 (0.43, 0.77) | 0.60 (0.42, 0.77) | 0.94 (0.89, 0.97) | 0.87 (0.79, 0.93) | 0.93 (0.88, 0.97) | 0.75 (0.62, 0.86) |
| Older | 30 | 0.63 (0.46, 0.78) | 0.72 (0.58, 0.84) | 0.82 (0.71, 0.90) | 0.73 (0.59, 0.84) | 0.90 (0.86, 0.95) | 0.85 (0.76, 0.92) |
| Normal weight | 19 | 0.61 (0.40, 0.80) | 0.63 (0.43, 0.81) | 0.92 (0.85, 0.96) | 0.77 (0.61, 0.89) | 0.95 (0.91, 0.98) | 0.65 (0.45, 0.83) |
| Overweight | 25 | 0.57 (0.37, 0.75) | 0.66 (0.49, 0.81) | 0.84 (0.74, 0.92) | 0.62 (0.44, 0.79) | 0.89 (0.82, 0.95) | 0.79 (0.65, 0.87) |
| Obese | 10 | 0.78 (0.55, 0.93) | 0.82 (0.61, 0.94) | 0.92 (0.81, 0.98) | 0.90 (0.76, 0.97) | 0.94 (0.86, 0.98) | 0.94 (0.85, 0.98) |
| Winter | 12 | 0.71 (0.47, 0.89) | 0.76 (0.54, 0.91) | 0.90 (0.79, 0.97) | 0.89 (0.77, 0.96) | 0.89 (0.76, 0.96) | 0.90 (0.78, 0.96) |
| Spring | 19 | 0.58 (0.36, 0.78) | 0.58 (0.36, 0.78) | 0.94 (0.88, 0.97) | 0.83 (0.71, 0.92) | 0.98 (0.95, 0.99) | 0.69 (0.49, 0.85) |
| Autumn | 34 | 0.58 (0.41, 0.73) | 0.65 (0.50, 0.78) | 0.85 (0.77, 0.91) | 0.68 (0.54, 0.81) | 0.91 (0.85, 0.95) | 0.80 (0.69, 0.88) |

LPA = light-intensity physical activity; MVPA = moderate-to-vigorous physical activity; SED = Sedentary time
Age (younger if age <50 years, older if age ≥50), BMI (normal weight if BMI<25.0, overweight if 25.0≤BMI<30.0, or obese if BMI≥30.0), and season of first data collection (winter, spring or autumn).

**Table 5:** Intraclass Correlation (ICC) Values for Different Subsets Based on Missing Data.

| Variables | All weeks<sup>a</sup> | Subset with no missing data<sup>b</sup> |
| --- | --- | --- |
| N | 71 | 65 |
| Time in bed (h/d) | 0.42 (0.33, 0.52) | 0.67 (0.61, 0.72) |
| Total Sleep (h/d) | 0.77 (0.72, 0.82) | 0.72 (0.66, 0.76) |
| SED (h/d) | 0.84 (0.80, 0.87) | 0.90 (0.88, 0.92) |
| SED30min (min/d) | 0.85 (0.81, 0.88) | 0.81 (0.77, 0.84) |
| LPA (min/d) | 0.91 (0.89, 0.93) | 0.92 (0.91, 0.93) |
| MVPA (min/d) | 0.97 (0.96, 0.97) | 0.77 (0.72, 0.80) |

*Notes:* ICCs are adjusted for age, BMI, and season when the data were collected.
LPA = light-intensity physical activity; MVPA = moderate-to-vigorous physical activity; SED = Sedentary time
<sup>a</sup> Have data at week 1 and either week 2, week 3, week 4, or all of them.
<sup>b</sup> Have complete data at week 1, week 2, week 3 and week 4.

**Table 6:** Intraclass Correlation Values (ICC) excluding the first day of measurement

| Variables | Model 1<sup>a</sup> | Model 2<sup>b</sup> |
| --- | --- | --- |
| Time in bed (h/d) | 0.71 (0.66, 0.76) | 0.83 (0.81, 0.84) |
| Total Sleep (h/d) | 0.74 (0.70, 0.78) | 0.86 (0.85, 0.87) |
| SED (h/d) | 0.91 (0.89, 0.92) | 0.94 (0.93, 0.94) |
| SED30min (min/d) | 0.85 (0.82, 0.88) | 0.89 (0.88, 0.90) |
| LPA (min/d) | 0.92 (0.91, 0.94) | 0.95 (0.95, 0.96) |
| MVPA (min/d) | 0.74 (0.68, 0.77) | 0.85 (0.84, 0.86) |

*Notes:* ICCs are adjusted for age, BMI, and season when the data were collected.
LPA = light-intensity physical activity; MVPA = moderate-to-vigorous physical activity; Period = two-week measurement period; SED = Sedentary time
<sup>a</sup> ICCs using 7-day assessment period and removing first day.
<sup>b</sup> ICCs using 14-day assessment period and removing first day.

## 4. Discussion

The main findings of the present study indicate that individuals with AC spend 12.2 [IQR 2.1] h/d in SED, nearly 60% do not meet the minimum amount of MVPA recommended by the WHO for people with chronic conditions,^18^ and 80% of participants do not reach the minimum 7 h/d of sleep recommended by the American Academy of Sleep Medicine for adults.^51^ This highlights that these behaviors might represent potential risk factors that compromise the cardiovascular health of patients with AC in the mid- and long-term.^52–54^ Another important finding indicates that device-measured PA and SED are stable across weeks, suggesting a high reproducibility of the measure using a 7-day assessment period. However, for sleep parameters, a 14-day measurement would be more accurate as the stability of the measure increased substantially compared to a 7-day assessment.

Individuals diagnosed with AC have the contraindication of participating in high-intensity exercise and competitive sports because they have been related with accelerated disease progression, greater risk of sudden cardiac arrest and death.^12,13,26,55^ Studies, such as one conducted in Veneto, a region of Italy, confirm the deleterious effect of competitive sports in inherited cardiomyopathies.^56^ However, how these recommendations lead to lifestyle changes that result in low PA levels and large period of SED in patients with AC is poorly understood, which is significant since PA is robustly associated with a decrease in both cardiovascular disease development and mortality.^55–57^ This study provides a detailed description of PA, SED, and sleep over the full 24-hour cycle from raw accelerometer data in a sample of well-characterized patients with AC from Spain. Overall, 12.2 h/d of the participants’ daily time was classified as SED and 59.2% of them did not meet a minimum of 150 min/week of MVPA as recommended by the WHO physical activity guidelines.

To our knowledge, this is the first study that uses accelerometers to assess SED and PA in patients with AC. Previous research has relied on self-reported assessments of regular exercise and competitive sports,^12,13,26–28^ therefore, daily activity patterns have not been ever quantified in this population. In addition, the use of devices instead of self-reported tools prevents the potential response bias, such as social desirability bias (i.e., overestimation of PA which may be greater in men and in people with lower education) or recall bias that leads to inaccurate memories of PA. ^12,13,26–28^ Accelerometers allow monitoring the full 24-h activity cycle and provide complete information on the activity patterns, including the sleep period.^58^ Our study confirms that patients with AC have an insufficient level of PA and spent most of the day in SED with a median of 12.2 hours per day. A previous study with a sample of older Spanish adults,^59^ using accelerometers to describe activity patterns, shows that people with an average age of 70 years spend a similar amount of excessive time in SED and more than three times the levels of MVPA present in our sample. Further, most SED was accumulated in bouts lasting <30min, as the median for SED bouts lasting ≥30 min was 61.3 minutes per day. The insufficient PA and sedentary lifestyle is in line with lifestyle patterns previously described in individuals with related cardiac disorders.^60–62^ No marked differences were generally observed in activity patterns when adjusting by socio-demographic factors and BMI; however, the group of participants aged 50 years and older accumulated a higher amount of SED in periods of at least 30 minutes and there was a trend to reduce MVPA, with fewer participants reaching the minimum recommendations for PA. Similarly, when stratifying by BMI, MVPA declined in later life and with increasing BMI, findings that are similar to those described in the general population.^63–65^ Regarding sleep parameters, the median sleep time was 6.4 h/d with 90% of sleep efficacy, results that reveal how most of the participants (80%) did not reach the 7 h/d of sleep recommended in adults by the American Academy of Sleep Medicine. Sleep data obtained with the accelerometer were homogeneous between groups, but when considering different clinical characteristics, a decrease in both total sleep time and sleep efficiency was observed in the ICD and desmosomal mutations groups. In this sense, available evidence also indicates that sleep disturbances are highly prevalent in patients with ICD.^66^

In this study, we also analyzed the stability of accelerometer-assessed PA and SED over 4 consecutive weeks. In general, given that a good stability (ICC **≥** 0.80) is thought to be achieved by PA protocols with a minimum of four consecutive days,^67,68^ the most common approach uses seven consecutive days protocol.^69^ To analyze the longitudinal changes in PA, it is important to know how much of the variation corresponds to random changes from week to week, and how much to actual change in the behavior. The ICCs obtained when analyzing the stability of the sleep, SED, and PA metrics over four consecutive weeks and using 7-day assessment period were moderate to excellent and were similar with or without adjustment for age, BMI, and season. However, a 14-day assessment period improved stability for sleep, SED and PA metrics, turning moderate stability into good stability. Therefore, a 7-day assessment period in patients with AC would be sufficient to achieve a good stability, which would provide further accurate information about activity patterns in patients with AC, nevertheless, for time in bed and sleep variables that tend to be more irregular in patients with cardiac conditions, a 14-day assessment period may increase the stability of the measure from moderate to good. In addition, when stratifying by socio-demographic and anthropometric characteristics, higher ICCs, mainly related to MVPA, are observed for older and more obese groups.

These differences have also been reported in studies analyzing PA in healthy adults, which is related to their lower participation in physical activities and more uniform and sedentary lifestyles.^34,70^

This study, analyzing the lifestyle activity patterns in people living with AC, highlights the significant prevalence of sedentary behaviors in this population and subsequently reduced levels of PA, especially MVPA. Therefore, possible hypotheses based on related cardiac conditions about the prevalence of sedentary patterns are confirmed by this study, possibly being related to current evidence suggesting that competitive sports or high-intensity exercise might accelerate disease progression.^9–13^ This clearly suggest that many patients with AC could obtain important health-related benefits from restricting SED and being more physically active, even if these activities are of light or moderate intensities. The extent to which device-measured PA, SED and sleep are related to the presence of classical cardiovascular risk factors or ischemic cardiovascular disease in AC is a relevant matter for future research.

This study has limitations. The study was observational in nature and therefore averts making inferences about causality. On the other hand, a comparison of activity patterns between basic socio-demographic characteristics did not show significant changes, which we assume a larger study sample for some of the measures would be needed. Despite these limitations, this is the first study to objectively measure PA patterns in individuals diagnosed with AC.

## 5. Conclusion

The findings of this study indicate that individuals with AC engage in large periods of SED, as well as insufficient PA and sleep. In fact, nearly 60% of participants do not meet the minimum amount of PA recommended by the WHO for people living with chronic diseases and 80% do not meet the sleep recommendations recommended by the American Academy of Sleep Medicine for adults. Device-measured PA and SED are stable across weeks, suggesting a high stability of the measure using a 7-day assessment period, however, for sleep parameters, a 14-day measurement would be needed to reach a similar stability. Future studies should focus on identifying potential barriers to PA among individuals with AC, and provide the basis for interventions aimed to promote PA and improve overall health.

## Data Availability

All data produced in the present work are contained in the manuscript

## Acknowledgments

This work was funded by the Consejería de Salud y Familias, Junta de Andalucía (Ref. PIER-0231-2019). This article is part of a Doctoral Thesis to be submitted by JR-M to the Clinical Medicine and Public Health Doctoral Program at the University of Granada (Spain).

## Authors’ Contributions

DR-G conducted the analyses and was responsible for writing the first manuscript draft and subsequent revisions; JR-M collected data, critically reviewed and edited the manuscript, and critically reviewed the analysis plan; JHM critically reviewed and edited the manuscript, and oversaw the data analyses; JAV-H collected data, contributed to the first manuscript draft and critically reviewed the manuscript; AR-S collected data and reviewed the manuscript; JJ-J conceptualized the study, collected data, revised the analysis plan, and critically reviewed and edited the manuscript; AS-M conceptualized the study, collected data, revised the analysis plan, and critically reviewed and edited the manuscript. All the authors contributed with important intellectual content, have read and approved the final version of the manuscript, and agree with the order of presentation of the authors.

---

**Supplementary Table 1.** Spearman Rank Order correlation values (comparison of week data)

| Variables | Week1 vs. Week 2 | Week1 vs. Week 3 | Week1 vs. Week 4 |
| --- | --- | --- | --- |
| Total Sleep (h/d) | 0.62* | 0.56* | 0.72* |
| SED (h/d) | 0.86* | 0.87* | 0.86* |
| LPA (min/d) | 0.94* | 0.94* | 0.92* |
| MVPA (min/d) | 0.85* | 0.88* | 0.81* |

\*P < .05

## References

1. Hutchinson MD. Arrhythmogenic right ventricular cardiomyopathy. Card Electrophysiol Clin Case Rev 2020:493–497.

2. Bazoukis G, Letsas KP, Thomopoulos C, et al. Predictors of Adverse Outcomes in Patients with Arrhythmogenic Right Ventricular Cardiomyopathy: A Meta-Analysis of Observational Studies. Cardiol Rev 2019;27(4):189–197.

3. Sawant AC, Bhonsale A, te Riele ASJM, et al. Exercise has a disproportionate role in the pathogenesis of arrhythmogenic right ventricular dysplasia/cardiomyopathy in patients without desmosomal mutations. J Am Heart Assoc 2014;3(6).

4. Corrado D, van Tintelen PJ, McKenna WJ, et al. Arrhythmogenic right ventricular cardiomyopathy: evaluation of the current diagnostic criteria and differential diagnosis. Eur Heart J 2020;41(14):1414–1429.

5. Hoorntje ET, Te Rijdt WP, James CA, et al. Arrhythmogenic cardiomyopathy: Pathology, genetics, and concepts in pathogenesis. Cardiovasc Res 2017;113(12):1521–1531.

6. Groeneweg JA, Bhonsale A, James CA, et al. Clinical Presentation, Long-Term Follow-Up, and Outcomes of 1001 Arrhythmogenic Right Ventricular Dysplasia/Cardiomyopathy Patients and Family Members. Circ Cardiovasc Genet 2015;8(3):437–446.

7. Bhonsale A, Groeneweg JA, James CA, et al. Impact of genotype on clinical course in arrhythmogenic right ventricular dysplasia/cardiomyopathy-associated mutation carriers. Eur Heart J 2015;36:847–855.

8. van der Voorn SM, te Riele ASJM, Basso C, Calkins H, Remme CA, van Veen TAB. Arrhythmogenic cardiomyopathy: Pathogenesis, pro-arrhythmic remodelling, and novel approaches for risk stratification and therapy. Cardiovasc Res 2020;116(9):1571–1584.

9. Miles C, Finocchiaro G, Papadakis M, et al. Sudden Death and Left Ventricular Involvement in Arrhythmogenic Cardiomyopathy. Circulation 2019;139(15):1786–1797.

10. Thiene G, Nava A, Corrado D, Rossi L, Pennelli N. Right Ventricular Cardiomyopathy and Sudden Death in Young People. N Engl J Med 1988;318(3):129–133.

11. Corrado D, Zorzi A. Sudden death in athletes. Int J Cardiol 2017;237:67–70.

12. Ruwald AC, Marcus F, Estes NAM, et al. Association of competitive and recreational sport participation with cardiac events in patients with arrhythmogenic right ventricular cardiomyopathy: results from the North American multidisciplinary study of arrhythmogenic right ventricular cardiomyopathy. Eur Heart J 2015;36(27):1735–1743.

13. Ruiz Salas A, Barrera Cordero A, Navarro-Arce I, et al. Impact of dynamic physical exercise on high-risk definite arrhythmogenic right ventricular cardiomyopathy. J Cardiovasc Electrophysiol 2018;29(11):1523–1529.

14. Bull FC, Al-Ansari SS, Biddle S, et al. World Health Organization 2020 guidelines on physical activity and sedentary behaviour. Br J Sports Med 2020;54(24):1451–1462.

15. Maron BJ, Chaitman BR, Ackerman MJ, et al. Recommendations for physical activity and recreational sports participation for young patients with genetic cardiovascular diseases. Circulation 2004;109(22):2807–2816.

16. Pelliccia A, Solberg EE, Papadakis M, et al. Recommendations for participation in competitive and leisure time sport in athletes with cardiomyopathies, myocarditis, and pericarditis: position statement of the Sport Cardiology Section of the European Association of Preventive Cardiology (EAPC). Eur Heart J 2019;40(1):19–33.

17. Pelliccia A, Sharma S, Gati S, et al. 2020 ESC Guidelines on sports cardiology and exercise in patients with cardiovascular disease. Eur Heart J 2021;42(1):17–96.

18. Reid H, Ridout AJ, Tomaz SA, Kelly P, Jones N. Benefits outweigh the risks: a consensus statement on the risks of physical activity for people living with long-term conditions. Br J Sport Med 2022;56:427–438.

19. Young DR, Hivert MF, Alhassan S, et al. Sedentary Behavior and Cardiovascular Morbidity and Mortality: A Science Advisory From the American Heart Association. Circulation 2016;134(13):e262–e279.

20. Genuardi M V., Ogilvie RP, Saand AR, et al. Association of Short Sleep Duration and Atrial Fibrillation. Chest 2019;156(3):544–552.

21. Li X, Zhou T, Ma H, et al. Healthy Sleep Patterns and Risk of Incident Arrhythmias. J Am Coll Cardiol 2021;78(12):1197–1207.

22. Badran M, Yassin BA, Fox N, Laher I, Ayas N. Epidemiology of Sleep Disturbances and Cardiovascular Consequences. Can J Cardiol 2015;31(7):873–879.

23. Jeong S-W, Kim S-H, Kang S-H, et al. Mortality reduction with physical activity in patients with and without cardiovascular disease. Eur Heart J 2019;40(43):3547–55.

24. Lear SA, Hu W, Rangarajan S, et al. The effect of physical activity on mortality and cardiovascular disease in 130 000 people from 17 high-income, middle-income, and low-income countries: the PURE study. Lancet 2017;390(10113):2643–2654.

25. Elliott AD, Linz D, Mishima R, et al. Association between physical activity and risk of incident arrhythmias in 402 406 individuals: evidence from the UK Biobank cohort. Eur Heart J 2020;41(15):1479–1486.

26. Pelliccia A, Corrado D, Bjørnstad HH, et al. Recommendations for participation in competitive sport and leisure-time physical activity in individuals with cardiomyopathies, myocarditis and pericarditis. Eur J Cardiovasc Prev Rehabil 2006;13(6):876–885.

27. James CA, Bhonsale A, Tichnell C, et al. Exercise increases age-related penetrance and arrhythmic risk in arrhythmogenic right ventricular dysplasia/cardiomyopathy-associated desmosomal mutation carriers. J Am Coll Cardiol 2013;62(14):1290–1297.

28. Lie ØH, Dejgaard LA, Saberniak J, et al. Harmful Effects of Exercise Intensity and Exercise Duration in Patients With Arrhythmogenic Cardiomyopathy. JACC Clin Electrophysiol 2018;4(6):744–753.

29. Shiraishi Y, Niimi N, Goda A, et al. Assessment of physical activity using waist-worn accelerometers in hospitalized heart failure patients and its relationship with kansas city cardiomyopathy questionnaire. J Clin Med 2021;10(18).

30. Klompstra L, Kyriakou M, Lambrinou E, et al. Measuring physical activity with activity monitors in patients with heart failure: from literature to practice. A position paper from the Committee on Exercise Physiology and Training of the Heart Failure Association of the European Society of Cardiology. Eur J Heart Fail 2021;23(1):83–91.

31. Berlin JE, Storti KL, Brach JS. Using Activity Monitors to Measure Physical Activity in Free-Living Conditions. Phys Ther 2006;86(8):1137–1145.

32. Fjeldsoe BS, Winkler EAH, Marshall AL, Eakin EG, Reeves MM. Active adults recall their physical activity differently to less active adults: test-retest reliability and validity of a physical activity survey. Health Promot J Austr 2013;24(1):26–31.

33. Dyrstad SM, Hansen BH, Holme IM, Anderssen SA. Comparison of self-reported versus accelerometer-measured physical activity. Med Sci Sports Exerc 2014;46(1):99–106.

34. Kelly P, Fitzsimons C, Baker G. Should we reframe how we think about physical activity and sedentary behaviour measurement? Validity and reliability reconsidered. Int J Behav Nutr Phys Act 2016;13(1):1–10.

35. Keadle SK, Shiroma EJ, Kamada M, Matthews CE, Harris TB, Lee IM. Reproducibility of Accelerometer-Assessed Physical Activity and Sedentary Time. Am J Prev Med 2017;52(4):541–548.

36. Marcus FI, McKenna WJ, Sherrill D, et al. Diagnosis of arrhythmogenic right ventricular cardiomyopathy/dysplasia: proposed modification of the Task Force Criteria. Eur Heart J 2010;31(7):806–814.

37. Corrado D, Perazzolo Marra M, Zorzi A, et al. Diagnosis of arrhythmogenic cardiomyopathy: The Padua criteria. Int J Cardiol 2020;319:106–114.

38. Perez De Isla L, Lennie V, Quezada M, et al. New generation dynamic, wireless and remote cardiac monitorization platform: a feasibility study. Int J Cardiol 2011;153(1):83–85.

39. Clarke CL, Taylor J, Crighton LJ, Goodbrand JA, McMurdo MET, Witham MD. Validation of the AX3 triaxial accelerometer in older functionally impaired people. Aging Clin Exp Res 2017;29(3):451–457.

40. Small S, Khalid S, Dhiman P, et al. Impact of Reduced Sampling Rate on Accelerometer-Based Physical Activity Monitoring and Machine Learning Activity Classification. J Meas Phys Behav 2021;4(4):298–310.

41. Migueles JH, Rowlands A V., Huber F, Sabia S, Hees VT van. GGIR: A Research Community–Driven Open Source R Package for Generating Physical Activity and Sleep Outcomes From Multi-Day Raw Accelerometer Data. J Meas Phys Behav 2019;2(3):188–196.

42. Van Hees VT, Fang Z, Langford J, et al. Autocalibration of accelerometer data for free-living physical activity assessment using local gravity and temperature: an evaluation on four continents. J Appl Physiol 2014;117(7):738–744.

43. van Hees VT, Gorzelniak L, Dean León EC, et al. Separating movement and gravity components in an acceleration signal and implications for the assessment of human daily physical activity. PLoS One 2013;8(4).

44. van Hees VT, Renström F, Wright A, et al. Estimation of daily energy expenditure in pregnant and non-pregnant women using a wrist-worn tri-axial accelerometer. PLoS One 2011;6(7).

45. Van Hees VT, Sabia S, Anderson KN, et al. A Novel, Open Access Method to Assess Sleep Duration Using a Wrist-Worn Accelerometer. PLoS One 2015;10(11).

46. van Hees VT, Sabia S, Jones SE, et al. Estimating sleep parameters using an accelerometer without sleep diary. Sci Rep 2018;8(1).

47. Hildebrand M, Van Hees VT, Hansen BH, Ekelund U. Age group comparability of raw accelerometer output from wrist- and hip-worn monitors. Med Sci Sports Exerc 2014;46(9):1816–1824.

48. Hildebrand M, Hansen BH, van Hees VT, Ekelund U. Evaluation of raw acceleration sedentary thresholds in children and adults. Scand J Med Sci Sports 2017;27(12):1814–1823.

49. Menai M, Van Hees VT, Elbaz A, Kivimaki M, Singh-Manoux A, Sabia S. Accelerometer assessed moderate-to-vigorous physical activity and successful ageing: results from the Whitehall II study. Sci Reports 2017;7(1):1–9.

50. Rosner B. Fundamentals of Biostatistics. 5th ed. Belmont, CA: Duxbury Press; 2005.

51. Watson NF, Badr MS, Belenky G, et al. Recommended Amount of Sleep for a Healthy Adult: A Joint Consensus Statement of the American Academy of Sleep Medicine and Sleep Research Society. Sleep 2015;38(6):843.

52. Lavie CJ, Ozemek C, Carbone S, Katzmarzyk PT, Blair SN. Sedentary Behavior, Exercise, and Cardiovascular Health. Circ Res 2019;124(5):799–815.

53. Sweeting J, Ingles J, Timperio A, Patterson J, Ball K, Semsarian C. Physical activity in hypertrophic cardiomyopathy: Prevalence of inactivity and perceived barriers. Open Hear 2016;3(2):1–9.

54. Daghlas I, Dashti HS, Lane J, et al. Sleep Duration and Myocardial Infarction. J Am Coll Cardiol 2019;74(10):1304–1314.

55. Koo DL, Nam H, Thomas RJ, Yun CH. Sleep Disturbances as a Risk Factor for Stroke. J Stroke 2018;20(1):12.

56. Ramakrishnan R, Doherty A, Smith-Byrne K, et al. Accelerometer measured physical activity and the incidence of cardiovascular disease: Evidence from the UK Biobank cohort study. PLOS Med 2021;18(1):e1003487.

57. Corrado D, Basso C, Pavei A, Michieli P, Schiavon M, Thiene G. Trends in Sudden Cardiovascular Death in Young Competitive Athletes After Implementation of a Preparticipation Screening Program. JAMA 2006;296(13):1593–1601.

58. Nystoriak MA, Bhatnagar A. Cardiovascular Effects and Benefits of Exercise. Front Cardiovasc Med 2018;5:135.

59. Rosenberger ME, Fulton JE, Buman MP, et al. The 24-Hour Activity Cycle: A New Paradigm for Physical Activity. Med Sci Sports Exerc 2019;51(3):454–64.

60. Cabanas-Sánchez V, Esteban-Cornejo I, Migueles JH, et al. Twenty four-hour activity cycle in older adults using wrist-worn accelerometers: The seniors-ENRICA-2 study. Scand J Med Sci Sports 2020;30(4):700–708.

61. Shiba S, Shiba A, Hatada A. Differences in Physical Activity between Patients with Peripheral Artery Disease and Healthy Subjects. J Aging Res 2020;2020.

62. Gerage AM, Correia M de A, de Oliveira PML, et al. Physical activity levels in peripheral artery disease patients. Arq Bras Cardiol 2019;113(3):410–416.

63. Hansen BH, Holme I, Anderssen SA, Kolle E. Patterns of Objectively Measured Physical Activity in Normal Weight, Overweight, and Obese Individuals (20-85 Years): A Cross-Sectional Study. PLoS One 2013;8(1):1–8.

64. Tudor-Locke C, Brashear MM, Johnson WD, Katzmarzyk PT. Accelerometer profiles of physical activity and inactivity in normal weight, overweight, and obese U.S. men and women. Int J Behav Nutr Phys Act 2010;7(1):1–11.

65. Smith L, Gardner B, Fisher A, Hamer M. Patterns and correlates of physical activity behaviour over 10 years in older adults: Prospective analyses from the English Longitudinal Study of Ageing. BMJ Open 2015;5(4):1–5.

66. Habibović M, Mudde L, Pedersen SS, Schoormans D, Widdershoven J, Denollet J. Sleep disturbance in patients with an implantable cardioverter defibrillator: Prevalence, predictors and impact on health status. Eur J Cardiovasc Nurs 2018;17(5):390–398.

67. Sasaki JE, Júnior JH, Meneguci J, et al. Number of days required for reliably estimating physical activity and sedentary behaviour from accelerometer data in older adults. J Sports Sci 2018;36(14):1572–1577.

68. Hart TL, Swartz AM, Cashin SE, Strath SJ. How many days of monitoring predict physical activity and sedentary behaviour in older adults? Int J Behav Nutr Phys Act 2011;8(1):1–7.

69. Migueles JH, Cadenas-Sanchez C, Ekelund U, et al. Accelerometer Data Collection and Processing Criteria to Assess Physical Activity and Other Outcomes: A Systematic Review and Practical Considerations. Sports Med 2017;47(9):1821.

70. Saint-Maurice PF, Sampson JN, Keadle SK, Willis EA, Troiano RP, Matthews CE. Reproducibility of Accelerometer and Posture-derived Measures of Physical Activity. Med Sci Sports Exerc 2020;52(4):876–883.

